# Systemic Inflammation and CT-Derived Coronary Plaque Characteristics Among Asymptomatic US Adults: The Miami Heart Study

**DOI:** 10.1101/2023.12.08.23299752

**Authors:** Shubham Lahan, Kobina Hagan, Miguel Cainzos-Achirica, Javier Valero-Elizondo, Shozab S. Ali, Lara Arias, Anshul Saxena, Theodore Feldman, Michael J. Blaha, Michael D. Shapiro, Ron Blankstein, Garima Sharma, Sadeer Al-Kindi, Svati Shah, Ricardo Cury, Matthew J. Budoff, Jonathan Fialkow, Khurram Nasir

## Abstract

**Objective:** We aimed to evaluate the association between inflammatory markers and coronary plaque features on coronary computed tomography angiography (CCTA) among asymptomatic individuals.

**Methods:** Baseline data from Miami Heart Study — an ongoing prospective community-based study of a primary prevention cohort from the Greater Miami Area without prior known CAD — were used for this cross-sectional analysis. Independent variables included high-sensitivity C-reactive protein (hsCRP; <2 vs ≥ 2mg/L) and Interleukin-6 (IL–6; in tertiles). The outcomes of interest were CCTA-based plaque findings: any plaque, CAC>0, CAC>100, maximal stenosis >50%, and high-risk plaque. Multivariable logistic regression models were constructed to evaluate the association between inflammatory markers and coronary plaque features.

**Results:** We evaluated 2,342 participants (50.4% men; mean age 53.4±6.7 years, 47% Hispanic, 43% non-Hispanic White, 8.3% diabetes, 56% hypertension, 22% on statin therapy). After adjusting for age, sex, and race/ethnicity, hsCRP ≥2 mg/L was associated with increased odds of having any plaque on CCTA [odds ratio (OR), 1.31 (95% confidence interval [CI], 1.09–1.58)] and stenosis ≥ 50% [OR, 1.69 (95% CI, 1.18–2.41)]. participants with IL–6 levels in 3^rd^ tertile were associated with higher odds of detecting any plaque [OR, 1.59 (95% CI, (1.27–1.99)], CAC >0 [OR, 1.34 (95% CI, 1.06–1.69)], ≥50% stenosis [OR, 2.41 (95% CI, 1.56–3.81)], and any high-risk plaque [OR, 2.41 (95% CI, 1.56–3.81)] Further adjustment for LDL, diabetes, hypertension, obesity, and tobacco use yielded nonsignificant associations.

**Conclusion:** Elevated levels of hsCRP and IL–6 are associated with the presence of coronary plaque and stenosis on CCTA when adjusted for demographics. However, these associations became nonsignificant after adjusting for additional cardiovascular risk factors. Our findings suggest a role of systemic inflammation as a mediator of the effect of cardiovascular risk factors on coronary plaque burden.

## 1. Introduction

Compelling experimental evidence in the last several decades shows the importance of inflammation in not only the initiation but also in the evolution and maturation process of atherosclerosis.^1^ The accumulation of low-density lipoprotein in the arterial intima induces the expression of adhesion molecules, chemokines, proinflammatory cytokines, and other mediators of inflammation in macrophages and vascular wall cells.^2^ This local inflammatory process results in a chronic cycle of plaque formation, vascular injury, inflammation, and repair with arterial calcium deposition.^3,4^ Several population-based studies have shown that individuals with chronic inflammatory diseases have an elevated risk of cardiovascular disease independent of traditional cardiovascular risk factors. Among asymptomatic individuals, biomarkers of the systemic inflammatory response such as high-sensitivity C-Reactive Protein (hsCRP) and interleukin-6 (IL-6) have shown predictive value for future cardiovascular events.^2,5–7^ The relationship between systemic inflammatory biomarkers and their relationship with the burden and evolution of subclinical atherosclerosis in the asymptomatic population remains understudied.

Evaluation of subclinical atherosclerosis incorporates various imaging modalities such as coronary artery calcium (CAC) and coronary computed tomography angiography (CCTA) to assess the plaque burden, features, and vulnerability to rupture. Although CAC has emerged as a reliable marker of subclinical atherosclerosis, it fails to capture the complete spectrum of plaque morphology and the maturation process.^8,9^ CCTA, on the contrary, is an excellent non-invasive modality for visualizing coronary anatomy and plaque morphology.^10^

In the current study, we sought to evaluate the association between biomarkers of systemic inflammation and subclinical coronary plaque features among asymptomatic adults using the Miami Heart (MiHeart) Study at Baptist Health South Florida.

## 2. Methods

### 2.1 Study design & Data sources

The Miami Heart study at Baptist Health South Florida is an ongoing, community based, prospective cohort study comprising individuals from the Greater Miami Area who are free from clinically established CVD. Briefly, the study evaluates the presence, features, and prognostic ability of diverse markers of subclinical coronary atherosclerotic disease in a cohort of middle-aged men and women free of CVD who underwent CCTA at baseline. The rationale, design, and further details about MiHeart have been described previously.^8^

### 2.2 Study variables

#### 2.2.1 Dependent variable

At baseline, all the study participants (N = 2,459) were subjected to a non-contrast cardiac-gated CT scan for CAC testing prior to a contrast-enhanced cardiac-gated CCTA. The scans were performed using the GE Revolution (GE Healthcare) scans®. Before the test, participants received oral and or intravenous beta-blocker (metoprolol) as needed to attain a heart rate of <75 beats per minute. Additionally, 0.4 mg of sublingual nitroglycerine was administered immediately before angiographic image acquisition. The radiation dose was estimated from the dose-length product times the conversion factor (0.014 mSv/mGy*cm). The assessment of 17-segment plaque presence/burden and classification was performed at a central core imaging lab.^8^

The primary outcomes of interest for this study included the following CCTA findings — any plaque (vs no plaque), CAC > 0 (vs CAC = 0), CAC > 100 (vs CAC ≤100), maximal stenosis ≥ 50% vs (< 50%), and any high-risk plaque feature (vs no high-risk feature). The presence of at least one of the following plaque features on CCTA was regarded as high-risk — spotty calcifications, low attenuation plaque, positive remodeling index >1.1, and napkin ring sign.

#### 2.2.2 Independent variable

Independent variables of interest were hsCRP and IL–6. These three markers of inflammation were assessed from blood sample tests conducted at study recruitment: hsCRP (mg/L), IL-6 (pg/mL), and homocysteine (umol/L). In line with previous literature, hsCRP was dichotomized with a cutoff value of 2 mg/L.^11,12^ IL–6, on the other hand, was categorized into tertiles.^13^

#### 2.2.3 Covariates

Besides age, sex, and race/ethnicity (non-Hispanic white, non-Hispanic black, non-hispanic Asian, non-hispanic others, non-hispanic/ >1 race, hispanic/ latino, and unknown/ not disclosed), other baseline characteristics relevant to the study included level of education (less than high school, high school diploma/GED equivalent, some college with no degree, bachelors, post-graduate, and unknown), annual income (<$25,000; 25,000 to 74,000; 75,000 to 149,000; ≥150,000; and not disclosed), basal metabolic index (BMI, kg/m^2^), smoking status (never, former, current), serum levels of LDL-cholesterol (mg/dL), high-density lipoprotein-cholesterol (HDL-C, mg/dL), and non-HDL-cholesterol (non-HDL-C, mg/dL), diabetes, hypertension, lipid-lowering medication use (any and statins), aspirin use, mean systolic and diastolic blood pressure (mmHg), and 10-year atherosclerotic cardiovascular disease (ASCVD) risk based on the pooled cohort equation.^14^

#### 2.3 Statistical Analyses

For modeling, hsCRP was dichotomized at a cut-off of 2 mg/L and IL–6 and homocysteine were divided in tertiles. Additionally, they were included as continuous variables in the analysis and the estimates were obtained per 10-unit increment in hsCRP and IL-6 levels. The baseline characteristics of our study population were also stratified by hsCRP and IL-6 categories. Descriptive statistics were used to summarize baseline socio-demographics characteristics and clinical comorbidities. Continuous variables were represented as mean ± standard deviation if normally distributed or median (interquartile range) if otherwise. Categorical data were represented as proportions.

For each outcome, multivariable logistic regression models were constructed and odds ratios (OR) with corresponding 95% confidence intervals (CI) were used to evaluate their independent associations with the inflammatory markers. For hsCRP, subgroup with values <2mg/L was used as the reference, while for IL–6 and homocysteine, the lowest tertile groups were used as the reference.

The following hierarchical regression models were used: Model 1 – unadjusted; Model 2 – adjusted for age, sex, race/ethnicity; Model 3 – adjusted for variables included in model 2 plus LDL-C, HDL-C, triglycerides, diabetes, systolic blood pressure, BMI, smoking status; Model 4 – in addition to variables adjusted for in model 3, model 4 was adjusted for antihypertensive medications, aspirin, and statin use. For all analyses, a two-sided p-value <0.05 was deemed statistically significant. All analyses were conducted in R v 4.0.5.

## 3. Results

### 3.1 Study population

Of the 2,459 participants enrolled in the MiHeart study, 100 individuals did not have complete CCTA data and were, thus, excluded from the present analysis. Additionally, 23 patients missing data on systemic inflammatory markers of interest were excluded from the analysis, yielding a final sample of 2,336 individuals, as illustrated in ***eFigure 1, Data Supplement***. Overall, the median age of our study population was 53.4 (48.1–58.9) years, of which 50.4% were men. The largest racial/ethnic groups were Hispanics (47%), followed by non-Hispanic whites (43.1%). Around 8.3% of the patients had underlying diabetes, 56.2% were hypertensive, and about 15% used statin medications. Obese participants comprised 32.6% of the cohort and 3% were found to be current smokers.

### 3.2 Distribution of CRP levels among study participants and their baseline characteristics

A total of 859 patients had elevated hsCRP levels with the majority being women (55%). They were also more likely to be hispanic. The age distribution among the high and low hsCRP groups was similar. Patients with high hsCRP levels were more likely to have obesity, with median BMI of 30.7 (27.5, 34.5) Kg/m^2^, compared with 26.6 (23.7, 29.1) Kg/m^2^ among their counterparts, as shown in ***Table S1, Data Supplement***. High hsCRP participants were also more likely to have hypertension, diabetes mellitus, and dyslipidemia compared with those in the low hsCRP group. However, they were less likely to be taking lipid-lowering medications including statins.

### 3.3 Distribution of IL–6 levels among study participants and their baseline characteristics

When stratified by IL–6 tertiles (***Table S2, Data Supplement*)**, a similar trend was observed as with CRP. Participants in the 3^rd^ tertile were more likely to be females, hispanic, and obese with a median BMI of 30.0 Kg/m^2^ compared to 26.3 and 27.6 Kg/m^2^ among the lower and middle tertiles, respectively. They were 1.5 times more likely to have hypertension and twice as more likely to have diabetes. The participants in the upper tertile of IL-6 also had significantly higher levels of non-HDL cholesterol, triglycerides, and apolipoprotein-B (p < 0.001 for all).

### 3.4 Coronary plaque, high-risk features, and inflammatory markers

**Tables 1 and 2** delineate the prevalence of coronary plaque and high-risk plaque features. Briefly, when comparing groups based on their inflammatory markers, we found no significant differences in the prevalence of CAC. However, we observed a higher prevalence of any plaque on CCTA among participants in the upper tertile of IL–6. Individuals with elevated hsCRP and IL–6 levels were 1.5- and 2-fols more likely to have ≥50% coronary stenosis, respectively. no significant differences were found in the prevalence of high-risk coronary features across the groups. Similar to CRP and IL-6, participants in the upper homocysteine tertile also had a greater prevalence of any plaque, ≥50% stenosis on CCTA in addition to CAC ≥100 Coronary plaque features according to homocysteine tertiles is shown in ***Table S2, Data Supplement***.

**Table 1.**
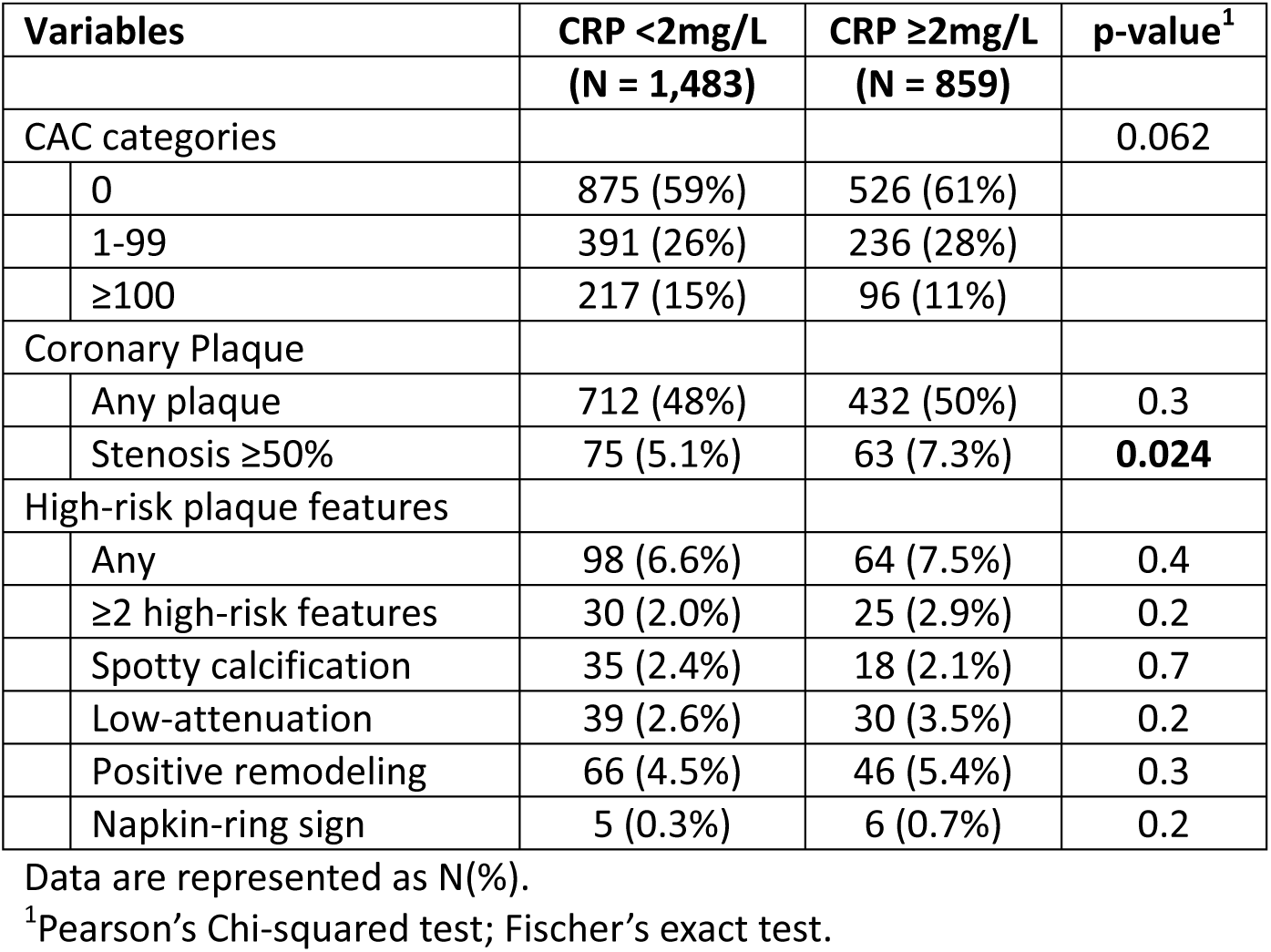
Coronary plaque characteristics of the study participants according to hsCRP levels.

| Variables |  | CRP <2mg/L<br>(N = 1,483) | CRP ≥2mg/L<br>(N = 859) | p-value<sup>1</sup> |
| --- | --- | --- | --- | --- |
| CAC categories |  |  |  | 0.062 |
|  | 0 | 875 (59%) | 526 (61%) |  |
|  | 1-99 | 391 (26%) | 236 (28%) |  |
|  | ≥100 | 217 (15%) | 96 (11%) |  |
| Coronary Plaque |  |  |  |  |
|  | Any plaque | 712 (48%) | 432 (50%) | 0.3 |
|  | Stenosis ≥50% | 75 (5.1%) | 63 (7.3%) | **0.024** |
| High-risk plaque features |  |  |  |  |
|  | Any | 98 (6.6%) | 64 (7.5%) | 0.4 |
|  | ≥2 high-risk features | 30 (2.0%) | 25 (2.9%) | 0.2 |
|  | Spotty calcification | 35 (2.4%) | 18 (2.1%) | 0.7 |
|  | Low-attenuation | 39 (2.6%) | 30 (3.5%) | 0.2 |
|  | Positive remodeling | 66 (4.5%) | 46 (5.4%) | 0.3 |
|  | Napkin-ring sign | 5 (0.3%) | 6 (0.7%) | 0.2 |

Data are represented as N(%).
<sup>1</sup>Pearson's Chi-squared test; Fischer's exact test.

### 3.3 Multivariable regression analyses

In the minimally adjusted model for age, sex, and race/ethnicity, hsCRP ≥2 mg/L was associated with higher odds of detection of any plaque on CCTA [odds ratio (OR), 1.33 (95% CI, 1.10–1.61)] and stenosis ≥ 50% [OR, 1.69 (95% CI, 1.11–2.41)] (**Table 3**). However, these associations were markedly attenuated when adjusted for additional CV risk factors and medication use. Similarly, when adjusted for age, sex, and race/ethnicity, participants with IL–6 levels in 3^rd^ tertile were associated with higher odds of detecting any plaque [OR, 1.59 (95% CI, (1.27–1.99)], CAC >0 [OR, 1.34 (95% CI, 1.06–1.69)], ≥50% stenosis [OR, 2.41 (95% CI, 1.56–3.81)], and any high-risk plaque [OR, 2.41 (95% CI, 1.56–3.81)], as shown in **Table 4**.

**Table 2.** Coronary plaque characteristics of the study participants according to IL-6 tertiles.

| Variables |  | IL-6 T1 | IL-6 T2 | IL-6 T3 | p-value |
| --- | --- | --- | --- | --- | --- |
|  |  | (N = 781) | (N = 781) | (N = 780) |  |
| CAC categories |  |  |  |  | 0.3 |
|  | 0 | 489 (63%) | 462 (59%) | 450 (58%) |  |
|  | 1-99 | 193 (25%) | 210 (27%) | 224 (29%) |  |
|  | ≥100 | 99 (13%) | 109 (14%) | 105 (13%) |  |
| Coronary plaque |  |  |  |  |  |
|  | Any plaque | 349 (45%) | 379 (49%) | 416 (53%) | **0.003** |
|  | Stenosis ≥50% | 31 (4.0%) | 37 (4.7%) | 70 (9.0%) | **<0.001** |
| High-risk plaque features |  |  |  |  |  |
|  | Any | 47 (6.0%) | 55 (7.0%) | 60 (7.7%) | 0.4 |
|  | ≥2 high-risk features | 11 (1.4%) | 23 (2.9%) | 21 (2.7%) | 0.1 |
|  | Spotty calcification | 16 (2.0%) | 23 (2.9%) | 14 (1.8%) | 0.3 |
|  | Low-attenuation | 15 (1.9%) | 25 (3.2%) | 29 (3.7%) | 0.1 |
|  | Positive remodeling | 31 (4.0%) | 38 (4.9%) | 43 (5.5%) | 0.4 |
|  | Napkin-ring sign | 2 (0.3%) | 4 (0.5%) | 5 (0.6%) | 0.5 |

Data are represented as N(%).
<sup>1</sup>Pearson's Chi-squared test; Fischer's exact test.

**Table 3.** Regression analysis for predicting CCTA plaque outcomes using hsCRP.

|  | Model 1 |  | Model 2 |  | Model 3 |  | Model 4 |  |
| --- | --- | --- | --- | --- | --- | --- | --- | --- |
|  | OR (95% CI) | p-value | OR (95% CI) | p-value | OR (95% CI) | p-value | OR (95% CI) | p-value |
| **Any Plaque** |  |  |  |  |  |  |  |  |
| hsCRP < 2mg/L | —Reference— |  |  |  |  |  |  |  |
| hsCRP ≥2mg/L | 1.10 (0.93, 1.30) | 0.287 | 1.33 (1.10, 1.61) | **0.003** | 0.92 (0.74, 1.15) | 0.454 | 0.98 (0.78, 1.23) | 0.875 |
| hsCRP per 10 mg/L | 1.13 (0.85, 1.51) | 0.387 | 1.58 (1.14, 2.21) | **0.007** | 1.16 (0.82, 1.64) | 0.411 | 1.23 (0.87, 1.75) | 0.248 |
| **CAC >0** |  |  |  |  |  |  |  |  |
| CRP < 2mg/L | —Reference— |  |  |  |  |  |  |  |
| CRP ≥2mg/L | 0.93 (0.78, 1.10) | 0.403 | 1.11 (0.92, 1.35) | 0.282 | 0.80 (0.64, 1.01) | 0.059 | 0.86 (0.68, 1.09) | 0.213 |
| hsCRP per 10 mg/L | 1.08 (0.81, 1.43) | 0.597 | 1.53 (1.09, 2.15) | **0.014** | 1.21 (0.85, 1.72) | 0.289 | 1.32 (0.92, 1.88) | 0.128 |
| **CAC ≥100<sup>§</sup>** |  |  |  |  |  |  |  |  |
| CRP < 2mg/L | —Reference— |  |  |  |  |  |  |  |
| CRP ≥2mg/L | -0.14 (-0.32, 0.05) | 0.142 | 0.04 (-0.13, 0.21) | 0.636 | -0.23 (-0.42, -0.05) | **0.013** | -0.16 (-0.34, 0.02) | 0.079 |
| hsCRP per 10 mg/L | 0.02 (-0.29, 0.32) | 0.919 | 0.29 (0.01, 0.57) | **0.045** | 0.06 (-0.22, 0.35) | 0.663 | 0.16 (-0.12, 0.43) | 0.274 |
| **Stenosis ≥50%** |  |  |  |  |  |  |  |  |
| CRP < 2mg/L | —Reference— |  |  |  |  |  |  |  |
| CRP ≥2mg/L | 1.49 (1.05, 2.10) | **0.025** | 1.69 (1.17, 2.41) | **0.004** | 1.01 (0.66, 1.53) | 0.977 | 1.05 (0.68, 1.59) | 0.834 |
| hsCRP per 10 mg/L | 1.02 (0.51, 1.70) | 0.96 | 1.18 (0.59, 2.03) | 0.592 | 0.66 (0.29, 1.24) | 0.249 | 0.71 (0.31, 1.35) | 0.351 |
| **Any high-risk plaque** |  |  |  |  |  |  |  |  |
| CRP < 2mg/L | —Reference— |  |  |  |  |  |  |  |
| CRP ≥2mg/L | 1.14 (0.82, 1.57) | 0.439 | 1.35 (0.95, 1.90) | 0.09 | 1.22 (0.82, 1.80) | 0.314 | 1.25 (0.84, 1.84) | 0.267 |
| hsCRP per 10 mg/L | 0.82 (0.38, 1.42) | 0.535 | 0.78 (0.30, 1.50) | 0.519 | 0.67 (0.26, 1.34) | 0.33 | 0.68 (0.26, 1.36) | 0.348 |

§Represents beta-coefficient with 95% CI. Model 1 – unadjusted; Model 2 – adjusted for age, sex, race/ethnicity; Model 3 – adjusted for age, sex, race/ethnicity, LDL cholesterol, HDL cholesterol, triglycerides, diabetes, systolic blood pressure, BMI, current smoker; Model – 4 adjusted for age, sex, race/ethnicity, LDL cholesterol, HDL cholesterol, triglycerides, diabetes, systolic blood pressure, BMI, current smoker, antihypertensive medications, aspirin therapy, statin, therapy.

**Table 4.** Regression analysis for predicting CCTA plaque outcomes using IL-6.

|  | Model 1 |  | Model 2 |  | Model 3 |  | Model 4 |  |
| --- | --- | --- | --- | --- | --- | --- | --- | --- |
|  | OR (95% CI) | p-value | OR (95% CI) | p-value | OR (95% CI) | p-value | OR (95% CI) | p-value |
| **Any Plaque** |  |  |  |  |  |  |  |  |
| T1 | —Reference— |  |  |  |  |  |  |  |
| T2 | 1.17 (0.96, 1.42) | 0.128 | 1.12 (0.89, 1.39) | 0.33 | 0.96 (0.76, 1.21) | 0.722 | 0.96 (0.76, 1.22) | 0.749 |
| T3 | 1.41 (1.16, 1.73) | **<0.001** | 1.59 (1.27, 1.99) | **<0.001** | 1.13 (0.88, 1.45) | 0.324 | 1.17 (0.90, 1.51) | 0.233 |
| IL-6 per 10 pg/mL | 0.94 (0.76, 1.12) | 0.501 | 0.97 (0.75, 1.18) | 0.767 | 0.96 (0.74, 1.17) | 0.707 | 0.94 (0.72, 1.17) | 0.655 |
| **CAC >0** |  |  |  |  |  |  |  |  |
| T1 | —Reference— |  |  |  |  |  |  |  |
| T2 | 1.14 (0.93, 1.39) | 0.216 | 1.05 (0.84, 1.32) | 0.673 | 0.93 (0.74, 1.18) | 0.571 | 0.94 (0.74, 1.20) | 0.646 |
| T3 | 1.23 (1.01, 1.51) | **0.043** | 1.34 (1.06, 1.69) | **0.013** | 0.99 (0.76, 1.27) | 0.921 | 1.02 (0.78, 1.33) | 0.888 |
| IL-6 per 10 pg/mL | 0.95 (0.77, 1.14) | 0.618 | 0.98 (0.75, 1.21) | 0.884 | 0.99 (0.75, 1.22) | 0.914 | 0.98 (0.73, 1.23) | 0.884 |
| **CAC ≥100<sup>§</sup>** |  |  |  |  |  |  |  |  |
| T1 | —Reference— |  |  |  |  |  |  |  |
| T2 | 0.11 (-0.11, 0.32) | 0.322 | 0.02 (-0.17, 0.21) | 0.843 | -0.07 (-0.27, 0.12) | 0.47 | -0.06 (-0.25, 0.13) | 0.542 |
| T3 | 0.19 (-0.02, 0.41) | 0.078 | 0.21 (0.02, 0.41) | **0.035** | -0.04 (-0.25, 0.17) | 0.71 | -0.02 (-0.22, 0.19) | 0.878 |
| IL-6 per 10 pg/mL | 0.00 (-0.19, 0.18) | 0.966 | 0.05 (-0.11, 0.21) | 0.562 | 0.05 (-0.11, 0.21) | 0.532 | 0.05 (-0.10, 0.20) | 0.523 |
| **Stenosis ≥50%** |  |  |  |  |  |  |  |  |
| T1 | —Reference— |  |  |  |  |  |  |  |
| T2 | 1.20 (0.74, 1.97) | 0.457 | 1.09 (0.66, 1.80) | 0.73 | 0.99 (0.59, 1.67) | 0.957 | 1.03 (0.61, 1.74) | 0.924 |
| T3 | 2.39 (1.56, 3.73) | **<0.001** | 2.41 (1.56, 3.81) | **<0.001** | 1.59 (0.97, 2.65) | 0.07 | 1.68 (1.02, 2.80) | **0.045** |
| IL-6 per 10 pg/mL | 1.03 (0.60, 1.29) | 0.866 | 1.07 (0.61, 1.37) | 0.72 | 1.10 (0.62, 1.42) | 0.591 | 1.10 (0.62, 1.42) | 0.593 |
| **Any high-risk plaque** |  |  |  |  |  |  |  |  |
| T1 | —Reference— |  |  |  |  |  |  |  |
| T2 | 1.18 (0.79, 1.77) | 0.413 | 1.09 (0.66, 1.80) | 0.73 | 0.99 (0.59, 1.67) | 0.957 | 1.03 (0.61, 1.74) | 0.924 |
| T3 | 1.30 (0.88, 1.94) | 0.192 | 2.41 (1.56, 3.81) | **<0.001** | 1.59 (0.97, 2.65) | 0.07 | 1.68 (1.02, 2.80) | **0.045** |
| IL-6 per 10 pg/mL | 1.00 (0.57, 1.27) | 0.993 | 1.01 (0.52, 1.34) | 0.965 | 1.05 (0.54, 1.38) | 0.823 | 1.03 (0.53, 1.37) | 0.892 |

§Represents beta-coefficient with 95% CI. Model 1 – unadjusted; Model 2 – adjusted for age, sex, race/ethnicity; Model 3 – adjusted for age, sex, race/ethnicity, LDL cholesterol, HDL cholesterol, triglycerides, diabetes, systolic blood pressure, BMI, current smoker; Model – 4 adjusted
for age, sex, race/ethnicity, LDL cholesterol, HDL cholesterol, triglycerides, diabetes, systolic blood pressure, BMI, current smoker, antihypertensive medications, aspirin therapy, statin, therapy.

We also explored the association of hsCRP and IL-6 as continuous variables with the coronary plaque outcomes. After accounting for sociodemographic factors, it was found that for every 10 mg/L increase in hsCRP levels, there were 58% higher odds of having any plaque [OR, 1.58 (95% CI, 1.14–2.21)] and 53% higher odds of CAC >0 [OR, 1.53 (95% CI, 1.09–2.15)] **(Table 3).** Nevertheless, there were no significant associations between CCTA outcomes and increase in IL-6 per 10 pg/mL (**Table 4**).

### 3.4 Causal Mediation Analysis

A causal mediation analysis was performed to evaluate the influence of hsCRP and IL–6 as the mediators of the effect of CV risk factors (diabetes, hypertension, LDL-C, Smoking, BMI) on the presence of any plaque (outcome). All risk factors except diabetes showed a significant direct effect on any plaque in the absence of mediators. However, the effect of CV risk factors through hsCRP and IL–6 (indirect effect) was non-significant (***Table S3 and S4, Data Supplement***).

## 4. Discussion

MiHeart is an ongoing, prospective cohort study enrolling individuals between the ages of 40 and 65 years free of underlying cardiovascular disease from the greater Miami area and who underwent a comprehensive evaluation of cardiovascular risk factors and various clinical measurements including but not limited to CCTA at baseline. In this study, we report results on 2,342 participants and the association between systemic inflammatory markers and coronary plaque features captured during their CCTA evaluation. Our analysis revealed that, upon adjustment for age, sex, and race/ethnicity, hsCRP levels of ≥2 mg/L were linked to increased odds of detecting any plaque and stenosis ≥50% on CCTA. Nevertheless, these associations lost significance when we further adjusted for additional cardiovascular risk factors and medication use. Similarly, participants whose IL-6 levels were in the 3rd tertile exhibited elevated odds of having any plaque, stenosis ≥50%, and any high-risk plaque, but these associations also became non-significant when the models were additionally adjusted for cardiovascular risk factors and medications.

Systemic inflammation is identified by a hyperactive immune system leading to an exaggerated immune response. Prior studies have shown elevated levels of hsCRP are commonly associated with high-risk plaque features such as low-attenuation plaque and napkin-ring sign on CCTA, which may promote plaque vulnerability and premature rupture of atherosclerotic plaque.^15,16^ This hypothesis is additionally supported by Pai et al, showing that elevated plasma levels of IL–6 and hsCRP translate into a high risk for coronary heart disease.^6^

Moreover, a recent study by Rubin and colleagues, comprising 1004 middle-aged asymptomatic South Korean patients, showed that hsCRP ≥2mg/L is associated with an odds ratio of 1.81 (95% CI, 1.19–2.74) and 1.66 (1.08–2.56) for detecting any plaque on CCTA when adjusted for sociodemographic variables and traditional cardiovascular risk factors, respcetively.^17^ Although the mean age of participants in both the studies is comparable (50 vs 53 years), compared to Rubin et al, our cohort with hsCRP ≥2mg/L had a higher prevalence any plaque (31% vs 50%). We, however, did not find a significant association when traditional cardiovascular risk factors were adjusted for. With that in mind, the current study holds significance because our results further build upon the correlation between coronary plaque features and elevated levels of IL–6, another valuable yet less commonly reported marker of systemic inflammation. Here, we additionally show that IL–6 levels in the 3^rd^ tertile are associated with significantly increased odds of detecting any plaque, CAC >0, CAC ≥100, ≥50% stenosis, and any high-risk plaque on CCTA after accounting for sociodemographic factors and that these associations lose significance when the model is further adjusted for traditional cardiovascular risk factors.

The results of our study warrant a cautious interpretion. The evidence pertaining to the role of inflammatory markers (chiefly hsCRP) in risk prediction still remains inconclusive.

Notably, while certain studies^17,18^ support the notion of hsCRP contributing to coronary plaque progression, others argue against it.^19,20^ A lack of independent association in our data may be due to a low statistical power. It is also plausible that systemic inflammation might be important in atherogenesis, but CRP may not be the optimal surrogate.

Our study has certain limitations. First, it is a cross-sectional study that did not aim to evaluate the causal mechanisms between systemic inflammation and coronary plaque features. Additionally, we had a limited study sample size to allow for precise estimation of the associations examined. However, the positive gradient observed between the biomarkers and subclinical atherosclerosis are consistent with prior studies on cardiovascular risk. Lastly, MiHeart has higher Hispanic population compared to the general U.S. population. While this limits the generalizability of our findings, it provides preliminary insights into the association between non-traditional risk factors and subclinical disease in a historically marginalized racial/ethnic group. Despite these limitations, this is one of the largest studies to evaluate the association between systemic inflammatory markers coronary plaque features in a community-based asymptomatic middle-aged population. In addition, we also explored the interplay between plaque and IL-6, which is further upstream in the inflammatory cascade.

## Conclusion

Elevated levels of hsCRP and IL–6 are associated with the presence of coronary plaque and stenosis on CCTA when adjusted for sociodemographic variables. These associations, however, are markedly attenuated when adjusted for additional cardiovascular risk factors, suggesting a potential modest role of systemic inflammation as a mediator of the effects of cardiovascular risk factors on coronary plaque burden.

## Data Availability

Data included in the manuscript will be made available upon request.

## Acknowledgements

None.

## References

1. Back M, Yurdagul A, Jr., Tabas I, Oorni K, Kovanen PT. Inflammation and its resolution in atherosclerosis: mediators and therapeutic opportunities. Nat Rev Cardiol. 2019;16(7):389–406.

2. Libby P, Ridker PM, Maseri A. Inflammation and atherosclerosis. Circulation. 2002;105(9):1135–1143.

3. El-Magadmi M, Bodill H, Ahmad Y, et al. Systemic lupus erythematosus: an independent risk factor for endothelial dysfunction in women. Circulation. 2004;110(4):399–404.

4. Herbrig K, Haensel S, Oelschlaegel U, Pistrosch F, Foerster S, Passauer J. Endothelial dysfunction in patients with rheumatoid arthritis is associated with a reduced number and impaired function of endothelial progenitor cells. Ann Rheum Dis. 2006;65(2):157–163.

5. Nasir K, Bittencourt MS, Blaha MJ, et al. Implications of Coronary Artery Calcium Testing Among Statin Candidates According to American College of Cardiology/American Heart Association Cholesterol Management Guidelines: MESA (Multi-Ethnic Study of Atherosclerosis). J Am Coll Cardiol. 2015;66(15):1657–1668.

6. Pai JK, Pischon T, Ma J, et al. Inflammatory markers and the risk of coronary heart disease in men and women. N Engl J Med. 2004;351(25):2599–2610.

7. Singh A, Collins BL, Gupta A, et al. Cardiovascular Risk and Statin Eligibility of Young Adults After an MI: Partners YOUNG-MI Registry. J Am Coll Cardiol. 2018;71(3):292–302.

8. Nasir K, Ziffer JA, Cainzos-Achirica M, et al. The Miami Heart Study (MiHeart) at Baptist Health South Florida, A prospective study of subclinical cardiovascular disease and emerging cardiovascular risk factors in asymptomatic young and middle-aged adults: The Miami Heart Study: Rationale and Design. Am J Prev Cardiol. 2021;7:100202.

9. Williams MC, Kwiecinski J, Doris M, et al. Low-Attenuation Noncalcified Plaque on Coronary Computed Tomography Angiography Predicts Myocardial Infarction: Results From the Multicenter SCOT-HEART Trial (Scottish Computed Tomography of the HEART). Circulation. 2020;141(18):1452–1462.

10. Parikh R, Patel A, Lu B, Senapati A, Mahmarian J, Chang SM. Cardiac Computed Tomography for Comprehensive Coronary Assessment: Beyond Diagnosis of Anatomic Stenosis. Methodist Debakey Cardiovasc J. 2020;16(2):77–85.

11. Arnett DK, Blumenthal RS, Albert MA, et al. 2019 ACC/AHA Guideline on the Primary Prevention of Cardiovascular Disease: A Report of the American College of Cardiology/American Heart Association Task Force on Clinical Practice Guidelines. Circulation. 2019;140(11):e596–e646.

12. Ridker PM, Danielson E, Fonseca FA, et al. Rosuvastatin to prevent vascular events in men and women with elevated C-reactive protein. N Engl J Med. 2008;359(21):2195–2207.

13. Cainzos-Achirica M, Enjuanes C, Greenland P, et al. The prognostic value of interleukin 6 in multiple chronic diseases and all-cause death: The Multi-Ethnic Study of Atherosclerosis (MESA). Atherosclerosis. 2018;278:217–225.

14. Goff DC, Jr., Lloyd-Jones DM, Bennett G, et al. 2013 ACC/AHA guideline on the assessment of cardiovascular risk: a report of the American College of Cardiology/American Heart Association Task Force on Practice Guidelines. Circulation. 2014;129(25 Suppl 2):S49–73.

15. Ait-Oufella H, Taleb S, Mallat Z, Tedgui A. Recent advances on the role of cytokines in atherosclerosis. Arterioscler Thromb Vasc Biol. 2011;31(5):969–979.

16. Tedgui A, Mallat Z. Cytokines in atherosclerosis: pathogenic and regulatory pathways. Physiol Rev. 2006;86(2):515–581.

17. Rubin J, Chang HJ, Nasir K, et al. Association between high-sensitivity C-reactive protein and coronary plaque subtypes assessed by 64-slice coronary computed tomography angiography in an asymptomatic population. Circ Cardiovasc Imaging. 2011;4(3):201–209.

18. Paffen E, DeMaat MP. C-reactive protein in atherosclerosis: A causal factor? Cardiovasc Res. 2006;71(1):30–39.

19. Reilly MP, Wolfe ML, Localio AR, Rader DJ, Study of Inherited Risk of Coronary A. C-reactive protein and coronary artery calcification: The Study of Inherited Risk of Coronary Atherosclerosis (SIRCA). Arterioscler Thromb Vasc Biol. 2003;23(10):1851–1856.

20. van der Meer IM, de Maat MP, Kiliaan AJ, van der Kuip DA, Hofman A, Witteman JC. The value of C-reactive protein in cardiovascular risk prediction: the Rotterdam Study. Arch Intern Med. 2003;163(11):1323–1328.

